# Utilizing a Variant Prioritization Approach to Explore Genetic Variants in Cervical Cancer Cases among Malawian Women

**DOI:** 10.64898/2026.10.23.26364490

**Authors:** Samuel Gwayi, Tamiwe Tomoka, Emile R. Chimusa, Yuri Fedoriw, Benjamin Kumwenda

## Abstract

**Introduction:** Cervical cancer (CC) represents a neoplastic transformation of the cervical epithelium. CC is the third most common cancer type in Malawi. In 2018, the estimated cervical cancer mortality rate was 51.5 deaths per 100,000 women per year among Malawian women. Although recent burden association and meta-analysis studies have identified candidate cervical cancer susceptibility variants in Malawian women, they did not establish the functional significance of individual variants.

**Aim:** This study aimed to identify germline genetic variants most likely to contribute to cervical cancer susceptibility among Malawian women.

**Methodology:** Whole blood samples were collected from 20 newly diagnosed cervical cancer patients at the Malawi National Cancer Treatment Centre between October and December 2023, and whole genome sequencing was performed. Variants were annotated via VEP and prioritized by ClinVar pathogenic status, Condel (SIFT and PolyPhen2) and minor allele frequency (MAF ≤ 0.1) in the 1000 Genomes project and GenomAD databases. Genes containing the prioritized variants were annotated using the DAVID tool.

**Results:** We identified five known pathogenic variants in the *ROS1* gene among Malawian cervical cancer patients. We also identified 51 moderate impact variants, 64.7% of which were located in known genes associated with CC, including *ADRB1, APOBEC3B, FAT1, IL12RB1, DST, EXO1* and *FGFRL1* genes.

**Discussion and Conclusion:** The presence of known pathogenic variants in the *ROS1* gene, which is associated with a high recurring rate of CC, as well as the identified moderate impact variants in other genes associated with CC, provide novel insights into the genetic landscape of cervical cancer in Malawi and highlight potentially functional variants that warrant further investigation.

## Introduction

Cervical cancer (CC) ranks as the fourth most common cancer by mortality among women worldwide, accounting for approximately 342,000 deaths in 2020. Over 90% of these fatalities occur in low- and middle-income countries, with the highest burdens concentrated in sub-Saharan Africa and Southeast Asia(1). Cervical cancer is classified as squamous cell carcinoma or adenocarcinoma. While adenocarcinoma incidence has increased recently, Squamous cell carcinoma is still more common and accounts for between 86% and 90% of all cervical cancer cases(2). Human papillomavirus (HPV) infection in the genital area accounts for more than 90% of cervical cancer cases (3). Upon entering the host cell, the human papillomavirus (HPV) integrates its genome into the host DNA. During this process, the viral E6 and E7 oncoproteins are expressed. These proteins degrade host tumour suppressor gene (P53) products and enhance the cellular response to growth factors. Since P53 gene products play crucial roles in the control of cell growth and development, their destruction, coupled with an enhanced cellular response to growth factors results in the abnormal development of host cells, leading to malignancy (4,5). While HPV infections account for most cervical cancer cases, HPV infections are usually transient in nature and are mostly cleared by the immune system through complicated mechanisms (6). Thus, recent studies have focused on other possible host-specific predisposing factors.

Through various genetic-based studies, approximately 112 Singe Nucleotide Polymorphisms(SNPs) linked to programmed cell death mechanisms and pathways related to the immune system functions have been associated with cervical cancer (7). The MHC is highly involved in the immune response as such, variations in the MHC have been associated with cervical cancer because the it plays a crucial role in immunity, especially by stimulates the development of immune cells that help fight against foreign antigens upon infection (8,9). Thus, while HPV infections can be effectively cleared by the immune system, variations in the MHC coupled with HPV infections can be associated with cervical cancer. While variations in genes related to the immune system have been associated with cervical cancer, other genes have been identified in the development of cervical cancer. These genes play a part in cell growth and differentiation, Deoxyribonucleic Acid (DNA) repair and apoptosis during the cell cycle (8). According to Ramachandran et al., 2022, there is a likelihood that many other variants associated with cervical cancer remain to be identified. This conclusion was drawn following the discovery of a novel SNP (rs225902), which is located close to the *FOXG1* and *PRKD1* genes, as a risk factor for cervical cancer. The role of the rs225902 was demonstrated through gene expression studies using cervical cancer cell lines (10). Another study conducted by Burk et al., 2017, with the overall aim of characterizing 228 primary cervical cancers, revealed that there were mutations in the *APOBEC, ARID1A, PTEN, SHKBP1, ERBB3, CASP8, HLA-A* and *TGFBR2* genes that are associated with cervical cancer. These discoveries further strengthened the association between genetic factors and cervical cancer, which could vary from one population to another.

While genetic-based studies conducted on cervical cancer have identified different gene variations associated with the development and progression of the disease, the findings are mostly limited to a specific population, and that only a few cervical cancer-related variants overlap between populations, suggesting that each population has a unique set of variants that predispose women to cervical cancer (12). This implies that for each population, especially those with a high cervical cancer incidence, a population-specific genetic study would be ideal.

In the Malawian population, the mortality rate is at 51.5 deaths per 100,000 women per year (13). Despite the CC mortality rate among Malawian women, only a limited number of studies have investigated the role of genetic variants in cervical cancer susceptibility within the population. Thus, current efforts are mostly directed towards HPV screening and vaccination against the disease. However, further exploration of the genetic basis of the disease may inform measures for both disease prevention and therapeutic management of advanced disease (14). A recent genetic study among Malawian cervical cancer cases identified a high burden of rare variants in genes related to lung, ovarian and osteosarcoma cancers suggesting shared genetic susceptibility across cancer types(15). Additionally, a meta-analysis identified a previously reported SNP in the *ITPR3* gene associated with cervical among Malawian women(16). However, these studies primarily provided statistical evidence of gene- and variant-level associations and did not establish the functional significance or biological plausibility of the identified variants. Hence, we employed a variant prioritization approach to identify potentially functional variants with the greatest likelihood of contributing to cervical cancer susceptibility, integrating functional annotation, predicted pathogenicity, population allele frequency and biological relevance to complement previous burden association and meta-analysis findings.

## Material and methods

Whole-genome sequencing data were generated from cervical cancer cases recruited in Malawi as previously reported. Briefly, genomic DNA was extracted from whole blood samples, sequenced using Illumina sequencing technology, and processed through GATK following best practices guidelines involving read alignment, variant calling, and quality control filtering. Detailed procedures for sample collection, sequencing, and initial variant processing have been reported previously(15,16). The resulting high-quality variant dataset was used for downstream variant prioritization analyses described in this study.

### Variant annotation and prioritization

In the current study, variants passing previous quality control filters were further analysed using a variant prioritization framework. Variant Effector Predictor (VEP) Ensembl version 108 was used to annotate the variants on the basis of their location and consequence on the coding sequence (17). High and moderate impact variants were processed further on the basis of their presence or absence in known genes associated with cervical cancer via ClinVar pathogenic predictions(18). Variants of Uncertain Significance (VUS) in ClinVar were further processed on the basis of the impact prediction of protein function via Consensus Deleteriousness (Condel) via SIFT and PolyPhen function predictors. Deleterious, probably or possibly damaging VUSs were further processed on the basis of minor allele frequency from the 1,000 Genomes Project database and Genome Aggregation Database (GenomAD) (https://gnomad.broadinstitute.org/news/2017-02-the-genome-aggregation-database/) to identify rare variants. These were variants with a population minor allele frequency (MAF) of less than 10% in a large population (19). After variants in known genes associated with CC were separated, all the pathogenic variants and rare variants were annotated on the basis of gene location in the Database for Annotation, Visualization, and Integrated Discovery (DAVID)(https://davidbioinformatics.nih.gov/tools.jsp) to assess for their association with CC(20).

## Results

### Cohort Demographics

Of the 20 Malawian participants, 12 (60%) were from the central region, 4 (20%) from the northern region, and the remaining 4 (20%) from the southern region. The average age of the participants was 46 years, with a median age of 50 years and an age range from 31 to65 years. Only 20% of the cases were diagnosed with Human Immuno-deficiency Virus(HIV).

### VEP and ClinVar variants classification

Of 17,443,881 variants that passed hard-filtering, 99.85% (17,418,319) were modifiers with no predicted effect on protein function. Of the remaining 25,562 variants, 61% (15,691) were classified as low-impact while the remaining 39% ( 9,871) comprised moderate- or high-impact variants, which were further characterized based on location as follows; moderate-impact coding variants accounted for 89% of the variants (8,791), while high-impact coding variants represented 4.2% (416). An additional 6.8% (664) of moderate- or high-impact variants were located in non-coding regions(Figure 1). Further filtering of the high and moderate impact variants based on known curated genes associated with CC as well as ClinVar pathogenic status revealed that 33 moderate-impact variants were located in genes associated with CC. Among these, 28 variants were non-pathogenic variants, while five variants were pathogenic variants located in *ROS1 gene* (**Figure 2**) and (**Table 1**). There were also 72 variants of uncertain significance (VUS), apart from 6,550 benign variants and 2,471 variants with no classification in the ClinVar database.

**Table 1.** Known pathogenic variants in ROS proto-oncogene 1, receptor tyrosine kinase. The number of cases with varying variants, the allele frequency in the 1000 Genomes Project(1000G_AF) and the genome aggregation database(GenomAD_AF), the variant reference SNP IDs and the nucleotide changes are included in the table.

| Variant ID | Variant Effect on Nucleotide | 1000G_AF | GenomAD_AF | No of Cases |
| --- | --- | --- | --- | --- |
| rs1998206 | ENST00000368507.8:<br>c.460A>C | 0.1677 | 0.1612 | 8 |
| rs12664076 | ENST00000368507.8:<br>c.5308G>C | 0.027 | 0.008698 | 1 |
| rs151330473 | ENST00000368507.8:<br>c.3388G>A | - | 4.78E-05 | 1 |
| rs143148482 | ENST00000368507.8:<br>c.3062A>T | 0.0036 | 0.0008396 | 1 |
| rs868736129 | ENST00000368507.8:<br>c.2126A>G | - | 1.19E-05 | 1 |

**Figure 1.**
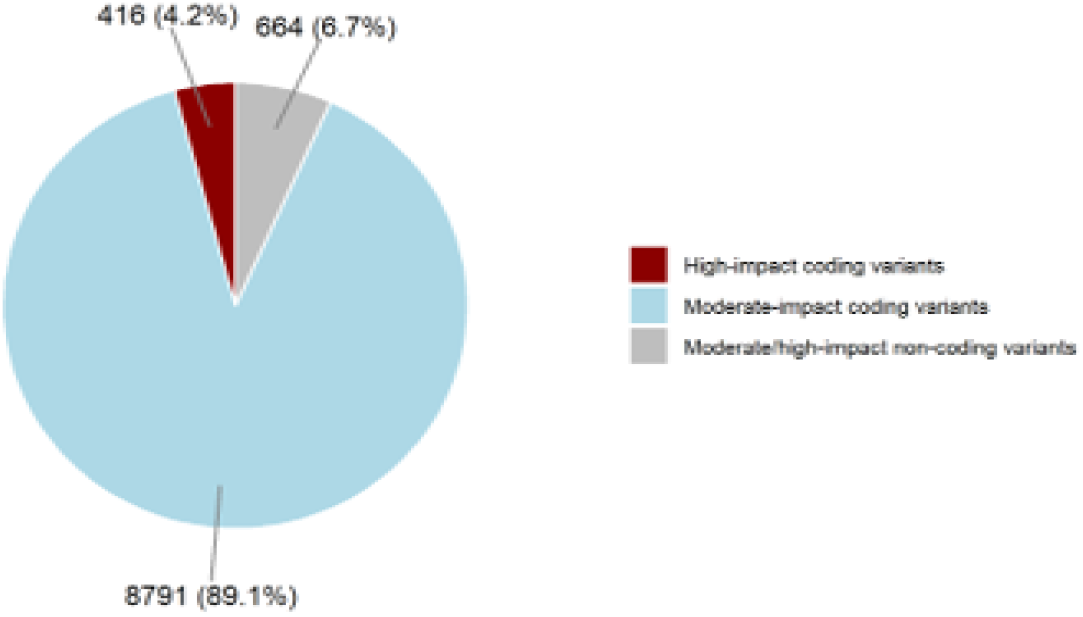
Impact-based classification of identified variants in Malawian cervical cancer patients via the use of an effector predictor. Moderate-impact variants cause considerable changes in genes or gene products that may affect phenotypes, whereas high-impact variants cause significant alterations in genes or proteins.

**Figure 2.**
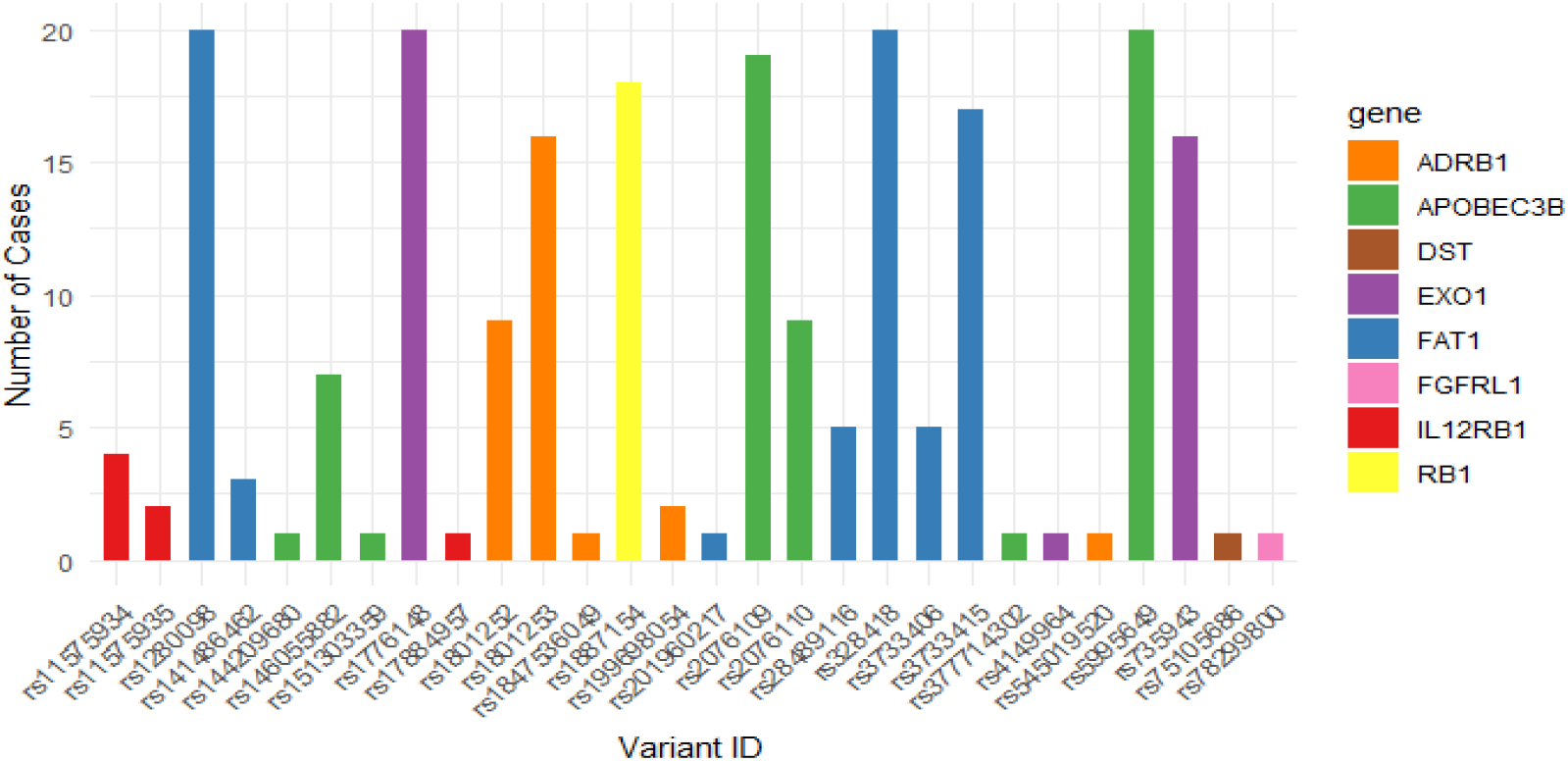
Known moderate impact ClinVar non-pathogenic variants present in genes associated with cervical cancer. The figure illustrates the number of Malawian cervical cancer patients carrying a specific variant in a particular gene. The genes listed are all curated known genes associated with cervical cancer.

### Classification Variants of Uncertain Significance

Processing of the VUS further on the basis of the SIFT and PolyPhen predictors revealed that only 48 variants were deleterious or damaging, while 16 variants were tolerated, and eight variants were of unknown impact. Among the deleterious and damaging variants, 18 variants were classified as rare VUS (MAF ≤ 0.1 in the 1000 Genomes and Genome Aggregation Database) as presented in the **Error! Reference source not found**.. However, on the basis of DAVID annotation, none of these deleterious rare variants were in genes directly linked to any cancer type.

## Discussion

This study investigated germline genetic factors among Malawian women that can be associated with cervical cancer. The high cervical cancer incidence and mortality rates in Malawi demand more robust and novel approaches to complement the available strategies in the prevention, control and management of cervical cancer cases. Our findings of pathogenic variants in the *ROS1* gene among Malawian cervical cancer patients as well as other variants with predicted impact on gene product in genes linked to CC, suggest a potential contribution of these variants to cervical cancer genetic variability and predisposition among Malawian women.

Among genes carrying known pathogenic variants, *ROS1* gene has many variants compared with the other identified genes, with the rs1998206 variant being present in 40% of cases. *ROS1* is characterized as a proto-oncogene(20). A recent study on the characterization of cervical cancer in the Japanese population suggested that *ROS1* gene mutations are present in recurring cases of cervical cancer (21,22). However, while our study identified pathogenic variants in gene, association studies would best demonstrate the role of the variants.

This study also identified moderate-impact variants in the *ADRB1, APOBEC3, FAT1, IL12RB1, DST, EXO1 and FGFRL1* genes among Malawian women with cervical cancer in addition to pathogenic variants in known genes associated with cervical cancer. In this study, 100% of the patients carried rs1280098, rs1776148, rs328418 or rs5995649 variants in the *APOBEC3B, EXO1* and *FAT1* genes. The *APOBEC3B* gene plays a major role in cancer regulation because of its involvement in cell growth and migration. This gene has been associated with the inhibition of programmed cell death through its interaction with tumour suppressor genes (23). High expression of *APOBEC3B* is associated with poor prognosis in cervical cancer patients because of its role in cancer metastasis and interaction with human papillomaviruses (HPV-16 and HPV-18), which are both oncogenes with a very high risk of causing cancer (24,25). On the other hand, single nucleotide polymorphisms (SNPs) in the *EXO1* gene, such as rs1047840 and rs10802996, have been associated with cervical cancer, especially among the Chinese population, with rs851797 being associated with poor cervical cancer disease prognosis (26,27). However, in this study, the Malawian patients carried different SNPs in this gene. The *FAT1* gene is also crucial for cancer control, as it has a role in tumour suppression. Mutations in this gene have been observed in many cancer types, including head and neck cancers and colorectal and breast cancers. Mutations reported in cervical cancer, suggest its role in disease development and progression (28).

There were 18 patients carrying a single variant, rs1887154, in the *RB1* gene, whereas 16 patients carried rs1801253 variants in the *ADRB1* gene in addition to other variants. Both *RB1* and *ADRB1* mutations have also been associated with cervical cancer. While *RB1* mutations are linked to cervical cancer susceptibility, ADRB1 mutations are associated with increased cervical cancer metastasis into the neurovascular system (29,30). *RB1* mutations have also been identified as either a causal or accelerating factor in other cancer types, including cancer of the retina, suggesting their significant role in many cancers (31). The *IL12RB1, DST and FGFRL1* genes also had mutations in only a single cervical cancer case for each gene. In a study on variation among genes related to immunity and cancer, it was observed that *IL12RB1* was among the genes that were mutated and thereby played a crucial role in the overall survival of cervical cancer patients, especially due to its link with lactic acid metabolism (32). The *DST* gene, on the other hand, has been identified as a new driver in cervical cancer and is among the top 10 most mutated genes in patients (33). The *FGFRL1* gene is one of the four membrane transduction receptor genes that play a role in signal transduction. A study on gene expression in cervical cancer revealed that, compared with squamous cell carcinoma, adenosquamous carcinoma had high expression of the *FGFRL1* gene.

Malawian cervical cancer patients also carry many rare variants that are predicted to be deleterious and damaging. These variants were present in genes not directly associated with any cancer type on the basis of DAVID annotation; however, further studies need to be performed to determine their contribution to cervical cancer.

### Study limitation

This study exclusively utilized samples from Malawian women, which may limit the generalizability of our findings. The genetic architecture of CC susceptibility may differ across populations; thus, our results may not be directly applicable to other ethnic or geographic groups.

Accordingly, future research should utilize larger and ethnically diverse cohorts to validate the role of these variants in CC causation within the Malawian population, as well as across broader populations.

## Conclusion

Our study identified pathogenic variants in the *ROS1* gene among Malawian women. The *ROS1* gene is a protooncogene that is associated with highly recurring cervical cancer. This study also revealed that moderate-impact variants in the *ADRB1, APOBEC3B, FAT1, IL12RB1, DST, EXO1* and *FGFRL1* genes are associated with cervical cancer. These findings provide novel insights into the genetic landscape of cervical cancer in Malawi and highlight potentially functional variants that warrant further investigation.

### Availability of data and materials

The datasets generated and/or analysed during the current study are available in the Genome Variation Map (GVM) in National Genomics Data Centre, Beijing Institute of Genomics, Chinese Academy of Sciences and China National Centre for Bioinformation repositor, under accession number GVM001125.

## Data Availability

All data produced in the present study are available upon reasonable request to the authors

## Ethics and conflicts of interest

This project was approved by the Malawi National Health Science Research Committee with approval number 4048, and the authors declare that they have no conflicts of interest.

## Acknowledgements

We are grateful to the National Health Institute (NIH) for their financial support through the Malawi Cancer Research Outcome Project (M-Corp), the Kamuzu University of Health Sciences (KUHeS) for their academic support during the project, and the staff members of the Malawi National Cancer Center, Kamuzu Central Hospital (KCH), UNC Lilongwe, KUHeS Molecular Laboratory and UNC Chapel Hill for their support during the project.

