## Supplemental Table S1 for "Utilizing a Variant Prioritization Approach to Explore Genetic Variants in Cervical Cancer Cases among Malawian Women"

**Supplementary table S1:** Rare variants with predicted deleterious and damaging effect on gene product present among Malawian cervical cancer cases. The variants were classified as rare variants in the 1000 genome and the genome aggregation database, and predicted to be deleterious and damaging. They were present in genes not linked to any cancer disease.

| **Gene** | **ID** | **Effect** | **1000G_AF** | **GenomAD_AF** | **No of Cases** |
|---|---|---|---|---|---|
| *DHX32* | rs143250708 | ENST00000284690.4:c.241G>A | 0.0002 | $7.98 \times 10^{-06}$ | 2 |
| *DNMBP* | rs116593720 | ENST00000324109.9:c.65C>T | 0.0002 | $9.55 \times 10^{-05}$ | 1 |
| *FRMPD1* | rs148192224 | ENST00000377765.8:c.770G>A | 0.0022 | 0.0008232 | 1 |
| *GDF7* | rs146317060 | ENST00000272224.5:c.1090G>C | 0.0004 | 0.0004601 | 1 |
| *HCAR1* | rs141141592 | ENST00000432564.3:c.418T>A | 0.0002 | $6.78 \times 10^{-05}$ | 1 |
| *KRT3* | rs200151587 | ENST00000417996.2:c.1208C>T | 0.0006 | 0.0001314 | 1 |
| *LBP* | rs2232613 | ENST00000217407.3:c.998C>T | 0.0551 | 0.0657 | 6 |
| *PKHD1L1* | rs77310972 | ENST00000378402.10:c.7715G>A | 0.0234 | 0.006034 | 7 |
| *PKHD1L1* | rs16879659 | ENST00000378402.10:c.10231A>G | 0.0615 | 0.0264 | 6 |
| *PKHD1L1* | rs114105023 | ENST00000378402.10:c.4084A>C | 0.0068 | 0.001566 | 2 |
| *PKHD1L1* | rs16879428 | ENST00000378402.10:c.1119G>C | 0.0162 | 0.003239 | 1 |
| *PKHD1L1* | rs187641517 | ENST00000378402.10:c.5614A>C | 0.0002 | $1.61 \times 10^{-05}$ | 1 |
| *TEX15* | rs139535218 | ENST00000256246.5:c.8033A>G | 0.0002 | 0.0001035 | 2 |
| *TEX15* | rs148337971 | ENST00000256246.5:c.6286T>G | 0.002 | 0.0003801 | 1 |
| *TEX15* | rs77311674 | ENST00000638951.1:c.626A>C | 0.004 | 0.0004389 | 1 |
| *TMEM132C* | rs559941011 | ENST00000435159.3:c.3286G>A | 0.0004 | $2.57 \times 10^{-05}$ | 2 |
| *UMODL1* | rs199705765 | ENST00000400424.6:c.86G>A | 0.0006 | 0.0001284 | 1 |
| *VWF* | rs145697622 | ENST00000261405.10:c.7150C>T | 0.0006 | 0.0002229 | 1 |
